# Indoor air pollutants and respiratory outcomes among residents of an informal urban setting in Uganda: a cross-sectional study

**DOI:** 10.1101/2022.07.28.22278151

**Authors:** Solomon T. Wafula, Aisha Nalugya, Hilbert Mendoza, Winnie K Kansiime, Tonny Ssekamatte, Abel Wilson Walekhwa, Richard K. Mugambe, Florian Walter, John C Ssempebwa, David Musoke

## Abstract

**Background:** Indoor air pollutants (IAP) such as particulate matter (PM) and carbon monoxide (CO) are a leading cause of acute respiratory symptoms, and long-term health impacts such as respiratory diseases, heart diseases and cancers. In Uganda, literature on the effects of IAP on respiratory outcomes in informal settlements is limited. This study investigated the association of selected IAPs and cooking fuels with respiratory symptoms among children and adults.

**Methods:** This study was conducted among 284 households in an informal settlement in Uganda from April to May 2022. Information on indoor air conditions, fuel type and adults reported the respiratory symptoms of their children as well as their respiratory symptoms within the previous 30 days. Same-day concentrations of PM less than 2.5 μm (PM_2.5_) and less than 10 μm(PM_10_) in diameter were monitored from 9 am to 2 pm using Temptop M2000c 2^nd^ edition particle sensor while CO was measured using a carbon monoxide meter. Robust Poisson regression was used to model the associations between indoor air conditions, fuel type and respiratory health outcomes.

**Results:** Approximately 94.7% of the households were using biomass fuels. Cough (66.2%), shortness of breath (33.5%) and phlegm (17.6%) were common respiratory symptoms in adults while in children, morning cough (80.0%), day or night cough (34.4%) and shortness of breath (26.5%) were reported. The median PM_2.5_, PM_10_ and CO levels were 49.5(IQR= 31.1, 86.2) µg/m^3^, 73.6(IQR= 47.3,130.5) µg/m^3^ and 7.7(IQR= 4.1,12.5) ppm respectively. Increase in humidity was associated with higher levels of PM_2.5_ (ß = 2.74, p =0.004) and PM_10_ (ß= 4.14, p =0.002) however temperature increases were associated with lower levels of PM_2.5_ (ß = -11.72, p =0.009) and PM_10_ (ß= -16.36, p =0.008) but higher CO levels (ß=2.24, p = 0.032). Use of less polluting fuels such as electricity and gas were associated with low levels of PM_2.5_ (ß= -31.36, 95%CI =-60.7 – -2.02) while home dampness (ß = 3.39, 95%CI =0.07 – 6.72) was associated with higher indoor CO levels. Dampness was associated with acute phlegm in adults (PR= 2.78, 95%CI =1.57 – 4.90) and outdoor cooking was found to be associated with lower shortness of breath risk (PR = 0.60,95%CI =0.40 – 0.91).

**Conclusion:** The prevalence of respiratory problems among adults and children was high. Poor indoor air conditions were associated with respiratory symptoms in adults and children. Efforts should be made to protect adults and children from the adverse effects of indoor air pollution.

## Introduction

Indoor air pollution (IAP) is a major environmental and public health challenge in low- and middle-income countries (LMICs) including Uganda(1). Approximately, 2 million people die prematurely per year from illnesses attributable to IAP from solid fuels such as charcoal and firewood (2). Sources of IAP include cooking and heating with solid fuels, burning candles or oil lamps, fuel-burning space heaters, tobacco smoke and indoor residual spraying against insect vectors. Of particular importance is inefficient and insufficiently vented cooking spaces and heating with biomass and coal on simple stoves in LMICs (3, 4). Burning these fuels in inefficient stoves results in poor combustion efficiency and high levels of emissions of health-damaging air pollutants including both fine and coarse particulate matter (PM), carbon monoxide (CO), nitrogen dioxide (NO_2_), sulphur dioxide (SO_2_), and a variety of volatile organic air pollutants (VOCs) (2).

The quantities emitted and relative composition of different emissions are determined by various factors including the type of fuel, humidity, dampness, stove type and the way the stove and fuel are used by the cook (5). There is growing evidence that IAP from use of biomass fuels can cause long-term health effects such as pneumonia, chronic obstructive pulmonary disease, heart disease, and lung cancer (6, 7). Additionally, a wide range of short-term health problems associated with poor indoor air quality (IAQ) have been documented including respiratory symptoms such as cough, phlegm, and wheezing (7, 8). Evidence suggests that women and children have a higher risk of adverse health effects compared to men (9, 10). These population groups (women and children) are more vulnerable due to prolonged exposure to biomass smoke during food preparation at home and spending much time indoors (11, 12).

Use of biomass fuels differs markedly by socioeconomic position with economically poor households more likely to use unimproved fuels (13, 14). In Kampala, the capital city of Uganda, the majority of dwellers use charcoal for cooking (15). Women especially in the informal settings are among the economically disadvantaged and live in overcrowded, poorly ventilated and have limited spaces for outdoor cooking and hence higher risk of health effects from biomass fuels. Women tend to cook from indoors or use poorly functioning stoves and firewood/charcoal; conditions that result in elevated levels of air pollutants such as PM, CO, and other VOCs (2, 16). Respiratory symptoms and diseases are among the adverse effects of exposure to elevated levels of these air pollutants. Previous studies have reported significant associations between respiratory problems and indoor sources of air pollutants, materials or activities, such as recent painting or new wall covering, volatile organic compounds (VOCs), gas appliances, biomass fuel use, and exposure to household smoking(17). However, there is limited data in sub-Saharan Africa on objectively measured concentrations of IAPs to enable a more robust assessment of the associations with respiratory effects. Moreover, although relying on sources of IAP may be a good proxy of exposure, objectively measured quantitative exposure assessments can be more useful for assessing the health effects of exposure. Our We aimed to investigate the association between selected indoor air pollutants, cooking fuels and objectively measured concentrations of air pollutants and respiratory symptoms among children and adults in an informal urban setting in Uganda.

## Methods

### Study setting

This study was conducted in Bwaise, an informal setting in Kawempe Division, Kampala, Uganda. Bwaise comprises one of the most densely populated slums on the outskirts of Kampala. These parishes are comprised of largely informal and substandard housing and small-scale businesses. It has a large population density, congested households with low socioeconomic status and high dependence on solid fuels (15). Therefore, we expected higher pollution levels in such a slum compared to other outdoor settings.

### Study design and population

This household-based cross-sectional survey used questionnaires and an observational checklist to collect data on self-reported respiratory symptoms including coughing, wheezing and phlegm among adults (aged 18 years and above) and children (aged 0 to 5 years), as well as individual risk factors in an informal settlement in Kampala, Uganda. Adults in the household who regularly do the cooking were primary respondents provided they had been residents in the area for at least three months. These adults also provided information on the demographics and respiratory health of all children (0 -59 months) under their care/household within the previous 30 days. Households were the study units.

### Sample size and sampling procedure

Using the Kish Leslie formulae for cross-sectional studies(18), we intended to sample 278 households using a prevalence of self-reported health effects (respiratory symptoms) of 19.4% (19), considering the power of 80% at a 5% level of significance and non-response rate of 15%.

As regards sampling, Bwaise slum was selected purposively because of poor housing and living conditions and over-reliance on solid/biomass fuels. Bwaise has three parishes; Bwaise 1, Bwaise II, and Bwaise III and participants were selected from each of these parishes as follows. Within each parish, one zone /village was selected randomly using the ballot method, and approximately 93 households were selected in each village using systematic sampling. The sampling interval for each village was obtained by dividing the number of households therein (according to records from the local council office) by the number of households needed (i.e., divided by 93). In each village, sampling started at the Chairperson’s place and then north direction then clockwise until the sample was obtained. A selected household was replaced by the next household if the original household had no eligible respondents or did not consent. In each selected household, one adult (mainly female) or another person generally involved in the cooking responded to the questionnaire. Information on respiratory symptoms among all children under 5 years of age was obtained from the adult in the selected household.

### Data collection and measurements

Experienced and trained data collectors with background training in Environmental Health Science administered the tools to eligible participants and responses were captured on mobile data collection app (Kobocollect; https://www.kobotoolbox.org/). Information collected included socio-demographic characteristics (age, marital status, income, education level), cooking fuel types (Biomass fuels(wood, charcoal,), improved fuels (electricity, gas, kerosene,), location of the cooking place (indoor vs outdoors), indoor air conditions (dampness, indoor plants, carpet use and indoor residual spraying for insects) and smoking within the household. Dampness was assessed through observation of the signs of mold growth and moist ground in indoor spaces. Data collection tools were developed following a thorough review of existing literature (20-23).

In addition, indoor levels of air pollutants of PM_2.5_, PM_10_, humidity and temperature were obtained. Same-day measurements were taken ten times at each household within 5 hours (twice per hour from 9 am to 2 pm) using the Low-cost particle sensor (Temptop M2000c 2^nd^ edition) which utilizes an optical particle counter. Temptop monitors were factory calibrated and therefore further laboratory calibration was not required for the duration of the study. CO levels were also measured ten times per household (twice per hour over 5 hours), using the AS8700A CO meter. These meters were placed 1 meter above the ground.

The primary outcome of the study was self-reported respiratory symptoms among adults and children (reported by parents/guardians) within the last 30 days. These symptoms included cough, phlegm, wheezing, shortness of breath and blocked/running nose. A secondary outcome was a composite outcome for overall prevalence of respiratory symptoms defined as any of the above respiratory symptoms (cough with phlegm, shortness of breath, wheezing and runny nose) reported in the 30 days before the survey.

### Statistical analysis

The continuous variables were described as median (IQR), while the categorical variables were shown as frequencies and percentages. Both continuous variables and categorical variables of air pollutants were used in regression analyses. The levels of air pollutants were also categorized as high and low levels using the median concentrations of air pollutants(24). We used robust least squares linear regression to assess the dependence of PM_2.5_, PM_10_ and CO on temperature, humidity, fuel types and other indoor conditions at the bivariate level. Two variables equal to 1/10 of PM2.5 and 1/10 of PM10 were used in the multivariable model as a predictor instead of original variables for better interpretation. We also performed bivariate and multivariable modified Poisson regression(25) producing prevalence ratios (PRs) and corresponding 95% confidence interval (95%CI) for the association of biomass use and particulate matter with adults and children’s respiratory symptoms; adjusting for age and gender. The data were analyzed using the statistical software Stata 14.0 version. All statistical tests were two-tailed, and statistical significance was assumed when a P value was <= 0.05.

### Ethical consideration

Ethical approval to conduct the study was obtained from Makerere University School of Public Health Higher Degrees Research and Ethics Committee (HDREC: Ref No. SPH-2021-99) and the Uganda National Council of Science and Technology (UNCST: Ref No. SS996ES). Confidentiality and anonymity were assured to the participants and their information. Written informed consent was obtained from each participant, children were not interviewed but their information was reported by parents/guardians.

## Results

### Background characteristics of the participants

More than three-quarters, 85.2% of the adult participants were females and 51.4% were aged below 30 years. The median age (IQR) was 29 years. Nearly half, 47.9% were married or living with a partner, 51.1% had attained a post-primary level of education and 68.6% were employed. More than half, 57.4% had a household monthly income that was between 50-150 US dollars, and 52.1% had stayed in their current area of residence for less than 5 years. Nearly a third, 31.7% of the participants cooked inside their living house. Of the 284 households, 187 had at least a child below the age of 5 years. The median age of the children was 24.0 months (IQR: 4,48). Of these, 52.6% were females and 72.2% were not in school (Table 1).

**Table 1:** Background characteristics of adult household participants.

| Characteristics | (N = 284) | % |
| --- | --- | --- |
| **Gender:** Female | 242 | 85.2 |
| **Age of respondents (in years)** |  |  |
| < 30 | 146 | 51.4 |
| 30 – 45 | 108 | 38.0 |
| > 45 | 30 | 10.6 |
| **Marital status** |  |  |
| Married or living with a partner | 136 | 47.9 |
| Separated | 43 | 15.1 |
| Single | 105 | 37.0 |
| **Education level** |  |  |
| No formal education | 25 | 8.8 |
| Primary | 114 | 40.1 |
| Post-primary | 145 | 51.1 |
| **Occupation** |  |  |
| Employed | 195 | 68.6 |
| Unemployed | 72 | 25.4 |
| Other | 17 | 6.0 |
| **Owner of the dwelling** | 26 | 9.2 |
| **Household monthly income (USD)** |  |  |
| < 50 | 77 | 27.1 |
| 50 – 150 | 163 | 57.4 |
| > 150 | 44 | 15.5 |
| **Duration of stay in the area of residence (years)** |  |  |
| < 5 | 148 | 52.1 |
| 5 – 10 | 57 | 20.1 |
| > 10 | 79 | 27.8 |

### Associations between cooking fuels, indoor conditions and air pollutant levels

Out of 284 households, 269 (94.7%) were using biomass fuels (wood and charcoal). Cooking from inside living spaces was reported in 90 households (31.7%), and indoor smoking in the previous 30 days was reported in 41 households (14.4%). The median PM_2.5_, PM_10_ and CO levels were 49.5 (IQR= 31.1,86.2) µg/m^3^, 73.6 (IQR = 47.3,130.5) µg/m^3^ and 7.70 (4.1,12.5) ppm respectively. Increase in humidity was associated with PM_2.5_ (ß = 2.74, 95%CI (0.87 – 4.61) and PM_10_ (ß= 4.14, 95%CI (1.48 – 6.80)) however increase in temperature was associated with lower PM_2.5_ (ß= -11.72, 95%CI (-20.49 – -2.94) and PM_10_ (ß = - 16.35, 95%CI (-28.4 – -4.30)). Use of less polluting fuel was associated with low levels of PM10. For CO, the levels increased with increases in temperature (ß=2.32, 95%CI (0.19 – 4.29)) and indoor home dampness (ß = 3.39, 95%CI (0.07 – 6.72)) (Table 2).

**Table 2:** Associations between cooking fuels, indoor conditions and air pollutant levels.

| Indoor air parameters | Summary statistic | PM2.5 Levels (Beta coefficients (95 % CI) | PM10 Levels (Beta coefficients (95 % CI) | CO Levels (Beta coefficients (95 % CI) |
| --- | --- | --- | --- | --- |
| **Meteorological parameters** |  |  |  |  |
| Humidity: Median (IQR) | 70.2 (66.9, 73.2) | **2.74 (0.87 – 4.61) \*\*** | **4.14 (1.48 – 6.80) \*\*** | - 0.15 (-0.67 – 0.37) |
| Temperature: Median (IQR) | 28.2 (27.2, 29.1) | **-11.72 (-20.49 – -2.94) \*\*** | **- 16.35 (-28.4 – -4.30) \*\*** | **2.32 (0.19 – 4.29) \*** |
| **Main Fuel type** |  |  |  |  |
| More polluting: n (%) | 269 (94.7) | 1 |  |  |
| Less polluting: n (%) | 15 (5.3) | -20.36 (-44.14 – 3.41) + | **-31.36 (-60.7 – -2.02) \*** | - 3.09 (-6.54 – 0.36) + |
| **Cooking from Outside** |  |  |  |  |
| No n (%) | 90 (31.7) | 1 | 1 | 1 |
| Yes n (%) | 194 (68.3) | -3.23 (-21.35 – 14.88) | - 6.99 (-31.28 – 17.31) | -1.64 (-5.08 – 1.80) |
| **Rearing pets n (%)** | 25 (8.8) | 21.14 (-12.44 – 54.74) | 30.35 (-17.12 – 77.81) | 0.26 (-3.96 – 4.49) |
| **Carpets in living room: n (%)** | 127 (44.7) | -4 .88 (-22.0 – 12.25) | -10.42 (-33.52 – 12.67) | 1.22 (-2.16 – 4.59) |
| **Home dampness (mould): n (%)** | 123 (43.3) | 12.94 (-4.68 – 30.57) | 19.50 (-4.17 – 43.17) + | **3.39 (0.07 – 6.72) \*** |
| **Smoking in the household: n (%)** | 41 (14.4) | 0.63 (-20.65 – 21.93) | 0.82 (-26.44 – 28.10) | 0.68 (-4.32 – 5.69) |

Note: \*p-value < 0.05, \*\*p-value <0.005, \*\*\*p-value<0.001

### Respiratory symptoms among adults and children

Of the 284 adults, 66.2% adults reported coughing, 41.9% reported a running nose, 33.5% reported shortness of breath, 17.6% reported phlegm and 14.8% reported wheezing. The majority of respondents (84.6%) reported having at least one of these respiratory symptoms in the previous 30 days. Among the 230 children, 80.0% had morning cough, 44.8% had a running nose, 34.4% reported day or night cough, 26.5% had shortness of breath, 20.0% had wheezing and 13.5% had phlegm during the previous 30 days.

### Associations between indoor air conditions and respiratory problems among adults

At multivariable analysis, marital status, occupation and household cooking place were associated with respiratory problems. The prevalence of wheezing was 2.48 times higher among single respondents compared to their married counterparts (PR=2.45, 95% CI = 1.18-5.22). Similarly, respondents whose cooking place was outside their living house had a 40% lower prevalence of shortness of breath compared to those whose cooking place was inside the living house (PR=0.60, 95% CI = 0.40-0.91). Respondents whose homes had dampness had a 181% higher prevalence of phlegm compared to those without (PR=2.81, 95% CI= 1.46-5.40). A ten-unit increase in PM _2.5_ levels was associated with a 1% higher risk of Cough among adults (PR = 1.01, 95%CI =1.01 – 1.02) (Table 4).

**Table 3:**
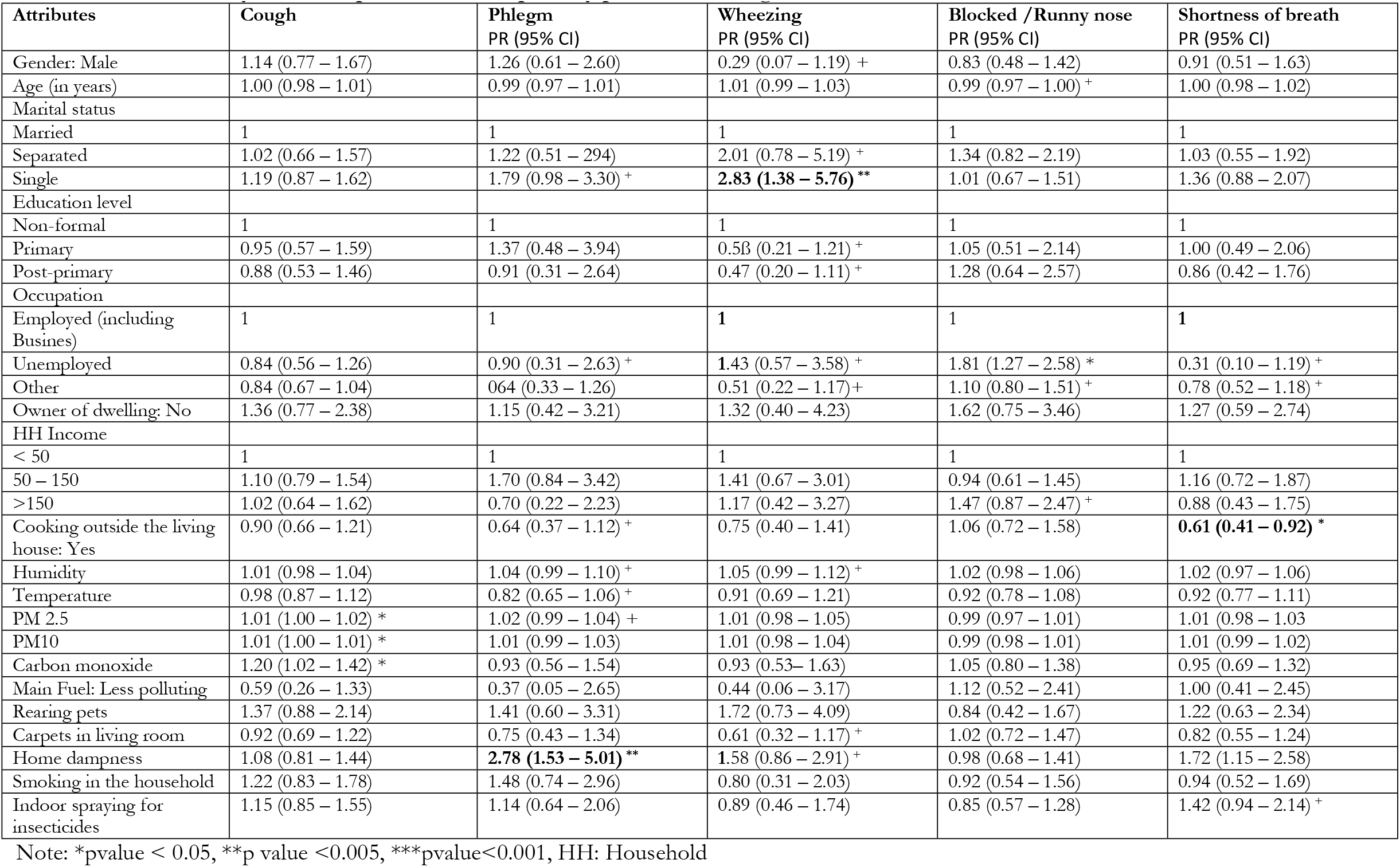
Bivariate analysis for the predictors of respiratory problems among adults.

| Attributes | Cough | Phlegm<br>PR (95% CI) | Wheezing<br>PR (95% CI) | Blocked /Runny nose<br>PR (95% CI) | Shortness of breath<br>PR (95% CI) |
| --- | --- | --- | --- | --- | --- |
| Gender: Male | 1.14 (0.77 – 1.67) | 1.26 (0.61 – 2.60) | 0.29 (0.07 – 1.19) + | 0.83 (0.48 – 1.42) | 0.91 (0.51 – 1.63) |
| Age (in years) | 1.00 (0.98 – 1.01) | 0.99 (0.97 – 1.01) | 1.01 (0.99 – 1.03) | 0.99 (0.97 – 1.00) + | 1.00 (0.98 – 1.02) |
| Marital status |  |  |  |  |  |
| Married | 1 | 1 | 1 | 1 | 1 |
| Separated | 1.02 (0.66 – 1.57) | 1.22 (0.51 – 2.94) | 2.01 (0.78 – 5.19) + | 1.34 (0.82 – 2.19) | 1.03 (0.55 – 1.92) |
| Single | 1.19 (0.87 – 1.62) | 1.79 (0.98 – 3.30) + | **2.83 (1.38 – 5.76) \*\*** | 1.01 (0.67 – 1.51) | 1.36 (0.88 – 2.07) |
| Education level |  |  |  |  |  |
| Non-formal | 1 | 1 | 1 | 1 | 1 |
| Primary | 0.95 (0.57 – 1.59) | 1.37 (0.48 – 3.94) | 0.53 (0.21 – 1.21) + | 1.05 (0.51 – 2.14) | 1.00 (0.49 – 2.06) |
| Post-primary | 0.88 (0.53 – 1.46) | 0.91 (0.31 – 2.64) | 0.47 (0.20 – 1.11) + | 1.28 (0.64 – 2.57) | 0.86 (0.42 – 1.76) |
| Occupation |  |  |  |  |  |
| Employed (including<br>Busines) | 1 | 1 | **1** | 1 | **1** |
| Unemployed | 0.84 (0.56 – 1.26) | 0.90 (0.31 – 2.63) + | **1.43 (0.57 – 3.58) +** | 1.81 (1.27 – 2.58) * | 0.31 (0.10 – 1.19) + |
| Other | 0.84 (0.67 – 1.04) | 0.64 (0.33 – 1.26) | 0.51 (0.22 – 1.17) + | 1.10 (0.80 – 1.51) + | 0.78 (0.52 – 1.18) + |
| Owner of dwelling: No | 1.36 (0.77 – 2.38) | 1.15 (0.42 – 3.21) | 1.32 (0.40 – 4.23) | 1.62 (0.75 – 3.46) | 1.27 (0.59 – 2.74) |
| HH Income |  |  |  |  |  |
| < 50 | 1 | 1 | 1 | 1 | 1 |
| 50 – 150 | 1.10 (0.79 – 1.54) | 1.70 (0.84 – 3.42) | 1.41 (0.67 – 3.01) | 0.94 (0.61 – 1.45) | 1.16 (0.72 – 1.87) |
| >150 | 1.02 (0.64 – 1.62) | 0.70 (0.22 – 2.23) | 1.17 (0.42 – 3.27) | 1.47 (0.87 – 2.47) + | 0.88 (0.43 – 1.75) |
| Cooking outside the living<br>house: Yes | 0.90 (0.66 – 1.21) | 0.64 (0.37 – 1.12) + | 0.75 (0.40 – 1.41) | 1.06 (0.72 – 1.58) | **0.61 (0.41 – 0.92) \*** |
| Humidity | 1.01 (0.98 – 1.04) | 1.04 (0.99 – 1.10) + | 1.05 (0.99 – 1.12) + | 1.02 (0.98 – 1.06) | 1.02 (0.97 – 1.06) |
| Temperature | 0.98 (0.87 – 1.12) | 0.82 (0.65 – 1.06) + | 0.91 (0.69 – 1.21) | 0.92 (0.78 – 1.08) | 0.92 (0.77 – 1.11) |
| PM 2.5 | 1.01 (1.00 – 1.02) * | 1.02 (0.99 – 1.04) + | 1.01 (0.98 – 1.05) | 0.99 (0.97 – 1.01) | 1.01 (0.98 – 1.03) |
| PM10 | 1.01 (1.00 – 1.01) * | 1.01 (0.99 – 1.03) | 1.01 (0.98 – 1.04) | 0.99 (0.98 – 1.01) | 1.01 (0.99 – 1.02) |
| Carbon monoxide | 1.20 (1.02 – 1.42) * | 0.93 (0.56 – 1.54) | 0.93 (0.53 – 1.63) | 1.05 (0.80 – 1.38) | 0.95 (0.69 – 1.32) |
| Main Fuel: Less polluting | 0.59 (0.26 – 1.33) | 0.37 (0.05 – 2.65) | 0.44 (0.06 – 3.17) | 1.12 (0.52 – 2.41) | 1.00 (0.41 – 2.45) |
| Rearing pets | 1.37 (0.88 – 2.14) | 1.41 (0.60 – 3.31) | 1.72 (0.73 – 4.09) | 0.84 (0.42 – 1.67) | 1.22 (0.63 – 2.34) |
| Carpets in living room | 0.92 (0.69 – 1.22) | 0.75 (0.43 – 1.34) | 0.61 (0.32 – 1.17) + | 1.02 (0.72 – 1.47) | 0.82 (0.55 – 1.24) |
| Home dampness | 1.08 (0.81 – 1.44) | **2.78 (1.53 – 5.01) \*\*** | **1.58 (0.86 – 2.91) +** | 0.98 (0.68 – 1.41) | 1.72 (1.15 – 2.58) |
| Smoking in the household | 1.22 (0.83 – 1.78) | 1.48 (0.74 – 2.96) | 0.80 (0.31 – 2.03) | 0.92 (0.54 – 1.56) | 0.94 (0.52 – 1.69) |
| Indoor spraying for<br>insecticides | 1.15 (0.85 – 1.55) | 1.14 (0.64 – 2.06) | 0.89 (0.46 – 1.74) | 0.85 (0.57 – 1.28) | 1.42 (0.94 – 2.14) + |

Note: \*pvalue < 0.05, \*\*p value <0.005, \*\*\*pvalue<0.001, HH: Household

**Table 4.** Independent predictors of respiratory problems among adult residents.

| Attributes | Cough<br>PR (95% CI) | Phlegm<br>PR (95% CI) | Wheezing<br>PR (95% CI) | Blocked /Runny<br>nose PR (95% CI) | Shortness of breath<br>PR (95% CI) |
| --- | --- | --- | --- | --- | --- |
| **Gender: Male** | 1.15 (0.93 – 1.43) | 1.66 (0.92 – 2.96)+ | 0.21 (0.03 – 1.39) + | 0.90 (0.52 – 1.58) | 0.91 (0.50 – 1.65) |
| **Age (in years)** | 0.99 (0.98 – 1.00) | 0.99 (0.96 – 1.01)+ | 1.00 (0.97 – 1.03) | 0.99 (0.97 – 1.00) + | 1.00 (0.98 – 1.01) |
| **Marital status** |  |  |  |  |  |
| Married |  | 1 | 1 |  |  |
| Separated |  | 1.56 (0.64 – 3.77) | 1.63 (0.56 – 4.77) |  |  |
| Single |  | 1.58 (0.91 – 2.73) + | **2.45 (1.18 – 5.22) \*** |  |  |
| **Education level** |  |  |  |  |  |
| No formal education | **1** |  | **1** |  |  |
| Primary | 0.85 (0.63 – 1.15) |  | 0.43 (0.19 – 0.72) * |  |  |
| Post-primary | 0.85 (0.63 – 1.14) |  | 0.44 (0.18 – 1.13) + |  |  |
| **Occupation** |  |  |  |  |  |
| Employed/ Business |  | 1 | **1** | 1 | 1 |
| Unemployed |  | 0.55 (0.19 – 1.63) + | 1.08 (0.39 – 1.52) + | 0.31 (0.08 – 1.14) | 0.65 (0.39 – 1.07) + |
| Other |  | 0.72 (0.37 – 1.41) | 0.67(0.31 – 1.52) | 0.76 (0.51 – 1.14) + | 0.29 (0.07 – 1.21) |
| **Household income** |  |  |  |  |  |
| < 50 |  |  |  | 1 |  |
| 50 – 150 |  |  |  | 0.95 (0.61 – 1.47) |  |
| >150 |  |  |  | 1.55 (0.92 – 2.63) |  |
| Cooking outside living house |  | 0.95(0.57 – 1.57) |  |  | **0.60 (0.40 – 0.91) \*** |
| Humidity |  | 1.01 (0.96 – 1.07) | 1.04 (0.97 – 1.11) |  |  |
| Carpets in living room |  |  | 0.61 (0.28 – 1.27) |  |  |
| Home dampness |  | **2.78 (1.57 – 4.90) \*** | 1.13 (0.61 – 2.09) |  |  |
| Indoor spraying for insecticides |  |  |  |  | 1.44 (0.95 – 2.19) + |
| PM2.5 | **1.01 (1.01 – 1.02) \*** | 1.01 (0.99 – 1.05) + | 0.99 (0.95 – 1.03) |  |  |
| High CO levels | 1.18 (0.99 – 1.41) | 0.92 (0.57 – 1.48) |  |  |  |

note: \*pvalue < 0.05, \*\*p value <0.005, \*\*\*pvalue<0.001

### Factors associated with parent-reported problems among children

At the bivariate level, the prevalence of phlegm among children in homes with dampness was 17.26 times (PR=17.26, 95% CI = 4.11-72.34) that among homes without dampness (Table 5).

**Table 5:** Bivariate analysis for the predictors of self-reported respiratory problems among children.

| Attributes | Morning Cough | Day or night cough | Phlegm | Wheezing | Blocked / Runny nose | Shortness of breath |
| --- | --- | --- | --- | --- | --- | --- |
| **Gender:** Male | 1.09 (0.81-1.45) | 0.68 (0.43 – 1.07) + | 1.04 (0.51 – 2.10) | 0.93 (0.52 – 1.67) | 0.86 (0.58 – 1.27) | 0.94 (0.57 – 1.56) |
| **Age (in years)** | 1.00 (0.99 – 1.00) | 1.00 (0.99 – 1.00) | 1.00 (0.99 – 1.01) | 1.00 (0.99 – 1.00) | 1.00 (0.99 – 1.00) + | 1.00 (0.99 – 1.00) |
| **Child Education** |  |  |  |  |  |  |
| Not in school | **1** | **1** | **1** | **1** | 1 | 1 |
| School | 1.05 (0.76 – 1.44) | 1.20 (0.75 – 1.93) | 1.24 (0.58 – 2.62) | 0.82 (0.41 – 1.61) | 0.92 (0.59 – 1.43) | 0.77 (0.42 – 1.40) |
| **Cooking place:**<br>**Outside** | 1.04 (0.76 – 1.42) | 1.07 (0.66 – 1.72) | 0.56 (0.28 – 1.14) + | 0.66 (0.37 – 1.19) + | 0.76 (0.51 – 1.14) + | 0.72 (0.43 – 1.20) |
| Humidity | 1.00 (0.97 – 1.03) | 1.00 (0.96 - 1.05) | 1.04 (0.97 – 1.11) | 1.05 (1.00 – 1.12) * | 0.99 (0.95 – 1.03) | 0.98 (0.93 – 1.03) |
| Temperature | 1.03 (0.91 – 1.18) | 1.14 (0.93 – 1.38) | 0.94 (0.69 – 1.29) | 0.88 (0.68 – 1.14) | 0.91 (0.77 – 1.09) | 1.17 (0.93 – 1.47) + |
| PM 2.5 | 1.02 (0.90 – 1.17) | 0.88 (0.61 – 1.26) | 1.89 (0.93 – 3.84) + | 0.90 (0.54 – 1.51) | 0.82 (0.61 – 1.09) | 1.13(0.73– 1.75) |
| PM10 | 1.00 (0.95 – 1.14) | 0.88 (0.61 – 1.26) | 1.89 (0.93 – 3.84) + | 0.90 (0.54 – 1.51) | 0.78 (0.59 – 1.05) | 1.06 (0.69 – 1.64) |
| High Carbon monoxide | 1.05 (0.92 - 1.20) | 0.91 (0.63 – 1.30) | 1.23 (0.63 – 2.39) | 1.05 (0.63 – 1.77) | 0.98 (0.73 1.30) | 1.19 (0.77– 1.84) |
| **Main Fuel:** More<br>polluting | 0.90 (0.44 – 1.82) | 1.56 (0.39 – 6.38) |  | 1.83 (0.25 – 13.29) | 1.36 (0.43 – 4.28) | 1.20 (0.29 – 4.92) |
| **Rearing pets** | 0.89 (0.57 – 1.41) | 1.24 (0.67 – 2.29) | 1.33 (0.51 – 3.47) | 1.69 (0.81 – 3.49) + | 0.58 (0.28 – 1.21) | 0.90 (0.41 – 1.97) |
| **Carpets in living**<br>**room** | 1.06 (0.79 – 1.43) | 1.09 (0.70 – 1.71) | 0.80 (0.38 – 1.66) | 1.02 (0.57 – 1.83) | 0.92 (0.62 – 1.36) | 1.14 (0.69 – 1.90) |
| **Home dampness** | 1.07 (0.80 – 1.43) | 1.42 (0.91 – 2.21) + | **17.26 (4.11 – 72.34)**<br>**\*\*\*** | 1.69 (0.94 – 3.04) + | 1.21 (0.82 – 1.79) | 1.31 (0.79 – 2.17) |
| **Smoking in the**<br>**household** | 0.98 (0.66 – 1.45) | 0.67 (0.34 – 1.34) | 0.77 (0.27 – 2.20) | 1.10 (0.51 – 2.35) | 1.18 (0.72 – 1.94) | 0.90 (0.44 – 1.83) |
| **Indoor spraying for**<br>**insecticides** | 1.04 (0.77 – 1.41) | 0.74 (0.45 – 1.21) | 3.03 (1.47 – 6.23) ** | 1.23 (0.68 – 2.22) | 2.04 (1.26 – 3.29) ** | 0.86 (0.50 – 1.48) |
| **Early life exposure to**<br>**SHS** |  |  |  |  |  |  |
| No exposure | 1 | 1 | 1 | **1** | 1 | 1 |
| Prenatal exposure or in the first 6 months | 0.96 (0.69 – 1.34) | 0.86 (0.51 – 1.45) | 0.31 (0.11 – 0.89) * | 1.03 (0.53 – 1.99) | 0.88 (0.56 – 1.39) | 0.68 (0.37 – 1.23) |
| Both | 0.87 (0.55 – 1.38) | 1.20 (0.66 – 2.19) | 0.64 (0.22 – 1.85) | 1.23 (0.55 – 2.74) | 1.19 (0.70 – 2.01) | 0.66 (0.30 – 1.48) |

Note: \*p value < 0.05, \*\*p value <0.005, \*\*\*p value<0.001

### Predictors of respiratory problems among children (Multivariate analysis)

At the multivariable level, the prevalence of phlegm among children in households that did indoor spraying of insecticides (PR=4.00, 95% CI = 1.69-9.45) was 4 times higher compared to that among households that did not. In addition, the prevalence of blocked/runny nose among children in households that did indoor spraying of insecticides (PR=1.99, 95% CI = 1.23-3.24) was 1.99 times that among households that did not (Table 6).

**Table 6:**
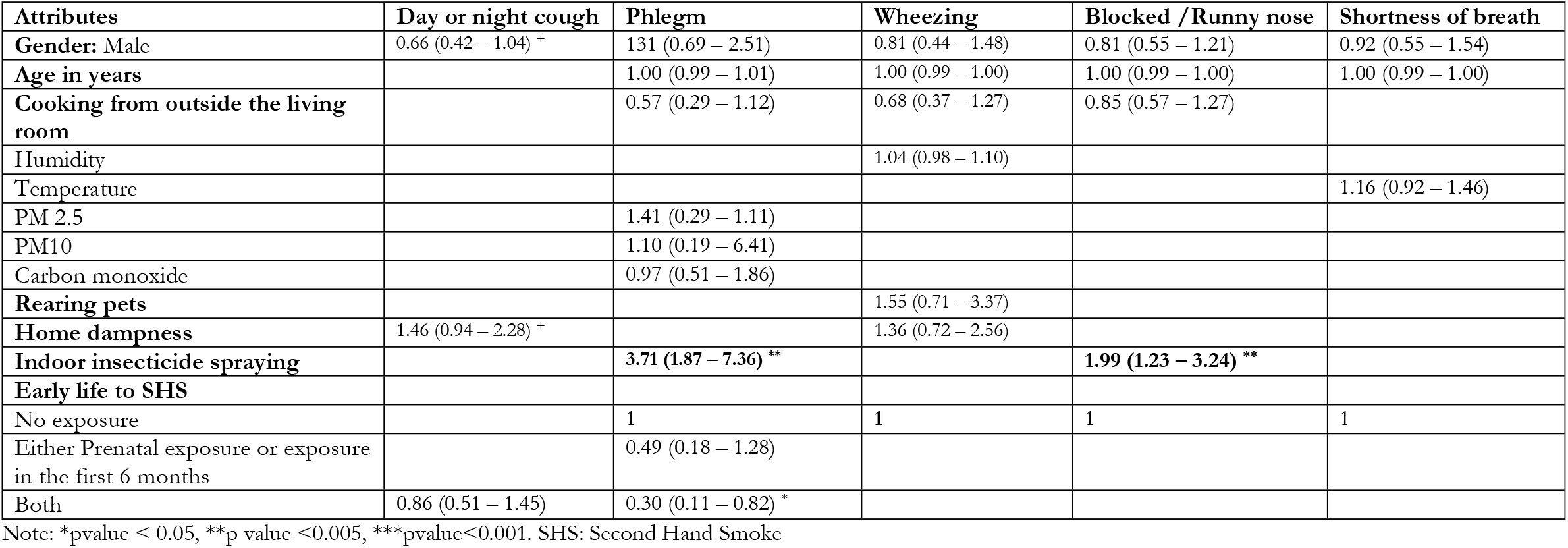
Independent Predictors of respiratory problems among children in Bwaise slum.

## Discussion

Understanding the effect of indoor air pollution on respiratory health is crucial because many people spend nearly two-thirds of their time at home (16). This is one of the first studies to investigate the association between indoor air quality conditions, biomass use, and respiratory symptoms among adults and children in informal settlements in Uganda. Average 5-hour same-day concentrations of PM_2.5_ and PM_10_ were high and significantly influenced by temperature, humidity and type of fuel used. Among adults, higher concentrations of PM_2.5_ were associated with an increased risk of cough. For children, we observed higher risks of phlegm and runny nose associated with indoor insecticide spraying.

The study found that the indoor levels of particulate matter (PM_2.5_ and PM_10_) increased with an increase in humidity. An increase in humidity leads to an increase in the moisture in the air, and as a result the particles become moist and increase in size, reducing dispersion of the particles, leading to an increase in the concentration of the PM_2.5_ and PM_10_ in the air (26, 27). In contrast, the study revealed that an increase in temperature was associated with lower concentrations of PM_2.5_ and PM_10_. Existing evidence shows that an increase in temperature leads to a decrease in humidity and an increase in air turbulence, and consequently a reduction in PM concentration (28). Our findings are similar to those in another cross-sectional study where a significant negative correlation was observed between PM_2.5_ and PM_10_ concentration and temperature (23). Use of less polluting fuel (gas and electricity) was associated with low levels of PM_2.5_ and PM_10_ compared to more polluting fuels such as charcoal and wood, which is consistent with other studies in India, South Africa, and Kenya (20, 22). This could be explained by the fact that the more polluting fuels (charcoal and wood) burn inefficiently and as a result emit higher levels of pollutants including PM_2.5_ and PM_10_(29). Our findings indicate a need for interventions that promote the use of clean fuels such as gas and electricity.

The study revealed a high prevalence (84.6%) of acute respiratory symptoms which included wheezing, cough and phlegm among adults in the last 30 days before the survey. Likewise, the majority of respondents reported their children had morning cough (80.0%), nearly half (44.8%) reported a runny nose, about a third (34.4%) reported day or night cough and 26.5% reported shortness of breath during the past month preceding the survey. Although self-reported, the high prevalence of respiratory symptoms highlights the significant burden among residents and requires efforts to reduce associated risk factors. Our study identified risk factors such as the location of the household cooking place, PM_2.5_, occupation, and marital status that were associated with respiratory problems in adults. In addition, the study indicated that the prevalence of Phlegm and blocked/runny nose in children was positively associated with indoor insecticide spraying. A 10 µg/m^3^ increase in PM_2.5_ levels was associated with a 1% higher risk of cough in adults which is consistent with published evidence (30-32). Indoor particulate matter concentration is related to a decrease in lung function and inflammation(33) and hence cough can be one of the manifestations of this effect. This underscores the need to reduce all activities that may encourage the deposition of particulate matter in indoor environments such as minimizing the use of biomass fuels and better ventilation. Whereas outdoor air quality was not assessed in this study, evidence elsewhere indicates that poor outdoor air quality could aggravate/increase the risk of respiratory symptoms in slum dwellers (34, 35). Further studies should also consider the role of outdoor pollution on respiratory morbidity.

The study also found that the risk of shortness of breath in adults whose cooking place was outside their living house was 40% lower than among those who were cooking inside their living spaces. This could be attributed to the fact that cooking inside the living house allows the accumulation of pollutants from the cooking fuels with limited ventilation, which affect respiratory health. When cooking occurs outside the house in an open area, the cooking smoke dissipates quickly, reducing exposure to individuals and hence lower risk of respiratory problems. Indeed, previous studies have reported a higher concentration of these pollutants in homes where cooking was done indoors (36). Our findings re-affirm those reported in another cross-sectional study in Thailand which indicated that cooking inside a home is associated with higher risk of respiratory symptoms, such as dyspnea (shortness of breath). Another cross-sectional study conducted in Ethiopia reported similar findings (37, 38). Therefore, there is a need to sensitize residents of informal settlements on the health risks associated with cooking indoors especially when using unclean fuels such as firewood and charcoal.

In our study, the prevalence of phlegm and blocked/runny nose was higher among children who resided in households that regularly conducted indoor insecticide spraying compared to those that did not. Indoor insecticide spraying is often done as a control measure for disease vectors including mosquitoes and other vectors. Indoor spraying, however, emits various indoor air pollutants including, hazardous air pollutants and volatile organic compounds which can settle on surfaces, furniture and counters. Overuse or incorrect use leads to the build-up of residues that may contribute to respiratory health problems when inhaled. This is consistent with findings from previous studies which revealed a positive relationship between respiratory symptoms in children under-five years and exposure to insecticides (39). Our findings indicate a need to emphasize and promote the use of non-chemical insect/pest control methods in informal settlements to reduce exposure to harmful chemical pollutants. Ensuring that enough time is allowed between spraying and occupation of the sprayed room can also ensure residual effect is minimized, hence lower risk of respiratory problems.

In this study, indoor house dampness was observed in nearly half of the participants’ homes which is not surprising since the slum setting is associated with frequent flooding and poor drainage. The prevalence of phlegm was higher among participants whose homes were characterised dampness and mould growth compared to those without which is consistent with previous literature(40, 41). Dampness creates an environment for mould spores to grow, which can trigger allergic reactions and respiratory problems. In addition, existing evidence indicates that dampness can initiate chemical or biological degradation of materials, and thus increase indoor concentration of pollutants which could increase risk of respiratory problems (21). Although there’s limited evidence of the association between residential dampness and phlegm in adults, Simoni, Lombardi (42) in a study conducted in Italy reported that dampness increased the risk of phlegm in children. Therefore, our findings indicate a need to implement strategies that minimize dampness in homes in informal settlements. The study also revealed that respondents who were not married had a higher prevalence of wheezing compared to their married counterparts. This could be because single individuals are often times more likely to exhibit harmful practices such as smoking (43, 44), which could increase their risk of developing respiratory problems. Further exploration of the effect of marital relationship status needs exploration to understand mechanisms by which it affects the risk of respiratory problems.

### Strengths and limitations

This is among the first studies to explore the associations between indoor air quality and respiratory health among adults and children in Uganda. Nevertheless, our study had some limitations regarding the design and interpretation of findings. This was a cross-sectional study in which causality between exposure to indoor air pollution and respiratory outcomes cannot be confirmed since temporality cannot be determined. Our measurement of air quality parameters was limited to 5 hours same day measurement and not 24 hours although we captured the most active period when most households would at least do some cooking and at least 10 measurements were made at each household throughout monitoring. There is a need to use prospective analytical study designs with longer duration of measurements to further examine the hypotheses and determine the predictors of respiratory problems among urban slum dwellers. In addition, the study was subject to recall bias as we relied on self-reports in the assessment of respiratory problems. Few households were using cleaner fuels, making it difficult to study the impact on respiratory outcomes.

## Conclusions

Our findings suggest that poor indoor air quality may have negatively affect the respiratory health of adults and children residing in urban informal settings. The poor indoor air quality may be majorly attributed to the reliance of most households on biomass fuels for cooking. Consequently, the study revealed indoor concentrations of PM_2.5_ and PM_10_ that were higher than WHO recommended exposure levels for most households. In the context of attainment of sustainable development goal 3, our study reemphasizes the need for interventions to reduce the risk of illnesses associated with indoor air pollution. Further prospective studies with longer duration measurement of pollutants are required to characterize the dose-response associations between the different types of household air pollutants and respiratory symptoms.

## Data Availability

Data are available from the corresponding author upon reasonable request

## Acknowledgements

We appreciate the research assistants and the participants for without which this study would not have been possible.

## Contributors

STW, DM, and JCS conceived and designed the study, STW, AN, AWW supervised data collection, STW, HM and AN analyzed the data and wrote the first draft of the manuscript. STW, AN, TS, WK,AWW, FW, RKM, DM, JC provided a critical review of the manuscript. All authors contributed to the interpretation of the results, review and editing of the manuscript and approved the final versional version of the manuscript for publication.

## Data availability statement

Data are available from the corresponding author upon reasonable request.

